# Evaluating the association between hybrid magnetic resonance positron emission tomography and cardiac-related outcomes in cardiac sarcoidosis

**DOI:** 10.1101/2022.01.13.22269230

**Authors:** Maria Giovanna Trivieri, Philip M. Robson, Vittoria Vergani, Gina LaRocca, Angelica M. Romero-Daza, Ronan Abgral, Nicolas A. Karakatsanis, Aditya Parikh, Christia Panagiota, Anna Palmisano, Louis DePalo, Helena L. Chang, Joseph H. Rothstein, Rima A. Fayad, Marc A. Miller, Valentin Fuster, Jagat Narula, Marc R. Dweck, Adam Jacobi, Maria Padilla, Jason C. Kovacic, Zahi A. Fayad

**Affiliations:** BioMedical Engineering and Imaging Institute, Icahn School of Medicine at Mount Sinai, New York, NY; Cardiovascular Institute, Icahn School of Medicine at Mount Sinai, New York, NY; School of Biomedical Engineering and Imaging Sciences, King’s College London, London, UK; Department of Cardiology, Hospital La Luz – Quirón Salud, Madrid, Spain; Department of Nuclear Medicine, University Hospital of Brest, European University of Brittany, EA3878 GETBO, Brest, France; Division of Radiopharmaceutical Sciences, Department of Radiology, Weill Cornell Medical College, New York, NY; Experimental Imaging Center, Department of Radiology, IRCCS San Raffaele Scientific Institute, Milan, Italy; Division of Pulmonary, Critical Care and Sleep Medicine, Icahn School of Medicine at Mount Sinai, New York, NY; International Center for Health Outcomes and Innovation Research, Department of Population Health Science and Policy, Icahn School of Medicine at Mount Sinai, New York, NY; Helmsley Electrophysiology Center, Icahn School of Medicine at Mount Sinai, New York, NY; British Heart Foundation Centre for Cardiovascular Science, University of Edinburgh, Edinburgh, UK; Department of Radiology, Icahn School of Medicine at Mount Sinai, New York, NY; Victor Chang Cardiac Research Institute, Darlinghurst, Australia and; St Vincent’s Clinical School, University of NSW, Australia

**Author notes:** Address for correspondence: Maria Giovanna Trivieri, MD, PhD One Gustave L. Levy Place, Box #1030 New York, NY 10029-6574. Authors contributed equally (Maria Giovanna Trivieri, Philip M. Robson).

**Keywords:** MR/PET, hybrid imaging, nuclear cardiology, cardiac sarcoidosis

## Abstract

**Objectives:** To evaluate an extended hybrid MR/PET imaging strategy in cardiac sarcoidosis (CS) employing qualitative and quantitative assessment of PET tracer uptake, and to evaluate its association with cardiac-related outcomes.

**Background:** Invasive endomyocardial biopsy is the gold standard to diagnose CS, but it has poor sensitivity due to the patchy distribution of disease. Imaging with hybrid late gadolinium enhancement (LGE) MR and ^18^F-fluorodexyglucose (^18^F-FDG) PET allows simultaneous assessment of myocardial injury and disease activity and has shown promise for improved diagnosis of active CS based on the combined positive imaging outcome, MR(+)PET(+).

**Methods:** 148 patients with suspected CS were enrolled for hybrid MR/PET imaging. Patients were classified based on presence/absence of LGE (MR+/MR-), presence/absence of ^18^F-FDG (PET+/PET-), and pattern of ^18^F-FDG uptake (focal/diffuse) into the following categories: MR(+)PET(+)_FOCAL_, MR(+)PET(+)_DIFFUSE_, MR(+)PET(-), MR(-)PET(+)_FOCAL_, MR(-)PET(+)_DIFFUSE_, MR(-)PET(-). Patients classified as MR(+)PET(+)_FOCAL_ were designated as having active CS [aCS(+)], while all others were considered as having inactive or absent CS and designated aCS(-). Quantitative values of standard uptake value (SUVmax), target-to-background ratio (TBRmax), target-to-normal-myocardium ratio (TNMRmax) and T2 were measured. Occurrence of a cardiac-related clinical outcome was defined as any of the following during the 6-month period after imaging: cardiac arrest, ventricular arrhythmia, complete heart block, need for cardiac resynchronization/defibrillator/pacemaker/monitoring device (CRT-D, ICD/WCD, or ILR). MR/PET imaging results were compared to the presence of the composite clinical outcome.

**Results:** Patients designated aCS(+) had more than 4-fold increased odds of meeting the clinical endpoint compared to aCS(-) (unadjusted odds ratio 4.8; 95% CI 2.0-11.4; p<0.001). TNMRmax achieved an area under the receiver operating characteristic curve of 0.90 for separating aCS(+) from aCS(-).

**Conclusions:** Hybrid MR/PET imaging with an extended image-based classification of CS was statistically associated with clinical outcomes in CS. TNMRmax had high sensitivity and excellent specificity for quantifying the imaging-based classification of active CS.

**Condensed Abstract:** Imaging with hybrid late gadolinium enhancement (LGE) MR and ^18^F-fluorodexyglucose (^18^F-FDG) PET allows simultaneous assessment of myocardial injury and disease activity and has shown promise for improved diagnosis of active cardiac sarcoidosis (CS). In this study, 148 patients with suspected CS were enrolled for hybrid MR/PET imaging. Patients were classified based on presence/absence of LGE (MR+/MR-), presence/absence of ^18^F-FDG (PET+/PET-), and pattern of ^18^F-FDG uptake (focal/diffuse). Patients classified as MR(+)PET(+)_FOCAL_ were designated as having active CS and, compared to patients with any other imaging pattern, they had more than 4-fold increased odds of cardiac-related outcome at 6 months from imaging.

## Introduction

Sarcoidosis is a rare multisystem, granulomatous disease that affects people of all ages and ethnic groups.(1) Among the several organs that can be infiltrated by sarcoid granulomas, the heart is an increasingly recognized site of disease involvement.(2) Clinically manifest cardiac sarcoidosis (CS) is present in 5-10% of patients,(3) but autopsy studies report cardiac involvement in at least 25% of cases.(4,5) Of note, active CS is associated with increased mortality independently of clinical manifestations.(2,5) For this reason, accurate diagnosis of subclinical disease could significantly impact on disease natural history and prognosis.

To date, diagnosis of CS is still lacking an effective gold standard. Histopathological diagnosis with endomyocardial biopsy is inherently insensitive, given the patchy distribution and intramyocardial localization of the disease.(3) A recent expert consensus statement suggested an integrated diagnostic approach using a combination of typical symptoms as well as findings on complementary cardiac imaging modalities that can identify pathological hallmarks of CS, namely inflammation, detected by ^18^F-fluorodeoxyglucose (^18^F-FDG) positron emission tomography (PET), and scar tissue, detected by cardiac magnetic resonance (MR) with late gadolinium enhancement (LGE) contrast.(6) In addition, both modalities have been proven to have an important prognostic role.(7,8)

Hybrid MR/PET imaging incorporates the advantages of both imaging techniques with a single scan and allows precise co-registration of LGE MR and ^18^F-FDG PET images. Accordingly, it appears to hold the greatest promise for the diagnosis of active CS, providing incremental information about both the pattern of injury and disease activity in a single scan.(9–11)

In our previous work,(9) we evaluated hybrid MR/PET imaging results of CS by assigning data to one of the four imaging categories: MR(+)PET(+), MR(+)PET(-), MR(-)PET(+), MR(-)PET(-). Of these, only MR(+)PET(+) was designated as positive for active cardiac sarcoidosis [aCS(+)]. All others were designated negative [aCS(-)]. The rationale for the joint designation was two-fold. Firstly, it is well-known that background physiological uptake of ^18^F-FDG in the myocardium can lead to false positive PET results; hence corroboration of disease was provided by the presence of scarring on LGE MR. Secondly, the presence of scarring on LGE MR can represent chronic or acute disease, and the concurrent finding of ^18^F-FDG uptake indicated active disease. This approach was still limited by the theoretical possibility of active inflammation occurring prior to manifest scarring, or of failed myocardial suppression of physiological uptake in patients with chronic scarring and inactive disease.

In our previous study,(9) we established that maximum target-to-normal-myocardium ratio (TNMRmax), a novel quantitative PET parameter to measure the ratio of PET tracer uptake at diseased and normal regions of the myocardium based on the presence of scarring on LGE MR, separated patients categorized as aCS(+) or aCS(-) by hybrid imaging. In addition, we noted that failed myocardial suppression often presented with intense and diffuse appearance of ^18^F-FDG uptake, as has been observed by others previously.(6,12)

In this study, we sought to confirm the role of hybrid MR/PET in a larger sample (148 patients) with known or suspected CS, using an extended imaging strategy to differentiate active from inactive myocardial disease and better identify false-positive ^18^F-FDG uptake. Importantly, we also sought to evaluate the potential prognostic capability of this imaging assessment by determining whether a designation of aCS(+) by imaging conferred a greater likelihood of a cardiac-related clinical outcome during follow-up of the same cohort.

## Methods

### Study population

148 patients with clinical suspicion of active CS due to established extra-cardiac involvement and/or clinical presentation suggestive of the disease were recruited at Mount Sinai Hospital, New York. This study was approved by our Institutional Review Board of the Icahn School of Medicine at Mount Sinai (GCO 01-1032) and all patients gave written informed consent. Exclusion criteria included: insulin-dependent diabetes mellitus, blood sugar >200 mg/dL prior to scanning, claustrophobia, pregnancy/nursing, presence of an implanted device contraindicated for MRI prior to the study, and impaired renal function (estimated glomerular filtration rate <40 ml/min/1.73 m^2^).

Additional clinical information was collected where available, including but not limited to: current treatment with corticosteroids and/or immunosuppresants; symptoms, including syncope, palpitations, dyspnea, and clinical heart failure; blood markers of inflammation and heart failure; and electrophysiological history, namely atrioventricular block, fascicular block, and bundle branch block.

### Patient follow-up

Patients were followed-up by chart review up to 6 months after their scan date to determine progression to a composite clinical endpoint including: cardiac-related death, aborted cardiac arrest, sustained ventricular arrhythmia, complete heart block, cardiac device requirement (pacemaker, cardiac resynchronization therapy device (CRT-D), implantable cardioverter defibrillator (ICD) or wearable cardioverter defibrillator (WCD), implantable loop recorder (ILR)). The decision to implant a cardiac device was made based on best clinical judgement and a combination of factors including clinical and imaging history and/or other indications such as inducible sustained ventricular arrhythmia at electrophysiological testing.

### Scanning protocol

Each patient underwent simultaneous cardiac MR with LGE and ^18^F-FDG PET cardiac imaging on a hybrid MR/PET system (Biograph-mMR, Siemens Healthineers).

#### PET imaging

PET imaging was performed as previously described.(9) Patients were required to fast for at least 4 hours, and to follow a low-carbohydrate high-fat diet for 24 hours prior to the study in concordance with published guidelines.(12) Patients were then injected intravenously with approximately 5 MBq/kg of ^18^F-FDG. PET data were subsequently acquired, in a single bed position centered on the heart, between 40-100 minutes post injection and reconstructed into a single static PET frame for both visual and quantitative PET analyses. PET image reconstruction employed an iterative ordinary Poisson ordered-subsets expectation-maximization algorithm with 21 subsets and 3 iterations incorporating point-spread-function resolution modeling, a 344×344×127 matrix and a 2-mm full-width-at-half-maximum Gaussian post-reconstruction filter. Attenuation correction for the body in the PET reconstruction was estimated using MR imaging. A recently proposed free breathing 3D radial gradient-echo acquisition was used to compensate for respiratory motion in the attenuation map.(13)

#### Cardiac MR sequences

Cardiac MR was performed simultaneously and included balanced steady-state free-precession (bSSFP) cine images (2-chamber, 4-chamber, and complete short axis stack) and T2-prepared bSSFP T2 mapping acquired in the basal, mid-ventricular and apical short-axis planes with three T2 preparation times (0, 25, 50 ms). Electrocardiograph-triggered, inversion-recovery fast gradient-echo LGE sequences were acquired 10 to 15 minutes after injection of 0.2 mmol/kg gadolinium contrast agent (Multihance, Bracco Imaging, Milan, Italy). Inversion times were optimized to null normal myocardium.

### Cardiac MR analysis

Left ventricular (LV) mass, biventricular volumes and ejection fractions were quantified from cine images by manually tracing the endocardial and epicardial contours on the short axis views in end-diastole and end-systole(14) using dedicated software (cvi42, Circle Cardiovascular Imaging, Calgary, Canada).

#### Late gadolinium enhancement

The presence and pattern of LV and right ventricular (RV) myocardial LGE (subendocardial, midwall, subepicardial, transmural) and whether this was characteristic of cardiac sarcoidosis were determined by 3 experienced operators (MGT, GL, AMRD). LGE was also quantified as a percentage of the total myocardial mass with dedicated software (cvi42, Circle Cardiovascular Imaging, Calgary, Canada).

#### T2

Basal, mid-ventricular and apical T2 maps were generated with dedicated software (cvi42, Circle Cardiovascular Imaging, Calgary, Canada), by two expert operators (VV, AMRD). A T2 value was extracted for each myocardial AHA segment, following the standard 16-segment AHA LV segmentation(15). The T2 value of the segment(s) including LGE was then selected. If no LGE was present, the average mid-ventricular T2 was used.

### PET analysis

#### Qualitative

Each scan was assessed systematically for extracardiac disease in the thoracic field. Analysis of myocardial ^18^F-FDG uptake on fused MR/PET datasets was performed using OsiriX-Lite software (OsiriX-imaging, Geneva, Switzerland). First, small adjustments were made to achieve accurate co-registration in 3 dimensions between the LGE MR and PET scans using anatomical markers.

#### Quantitative

PET quantification was performed on fused short axis LGE MR and ^18^F-FDG PET datasets by an experienced nuclear medicine physician (RA). Three different measures were recorded, as explained in our previous work:(9) maximum standard uptake values (SUVmax); maximum tissue-to-background ratio (TBRmax), where myocardial SUVmax was corrected for blood pool mean activity measured in the right ventricle; maximum target-to-normal-myocardium ratio (TNMRmax), where myocardial SUVmax was corrected for tissue mean activity measured in a contralateral myocardial segment without LGE. Maximum values were used in preference to mean values due to the difficulties in drawing consistent regions of interest.

### Hybrid MR/PET image analysis

#### Qualitative analysis to assign category

Co-registered short axis hybrid LGE MR and ^18^F-FDG PET images were then assessed and patients categorized into the following four groups: MR(+)PET(+), MR(+)PET(-), MR(-)PET(+), MR(-)PET(-), similarly to our previous study.(9) The PET(+) groups were further subdivided according to the characteristic appearance of the ^18^F-FDG PET signal, resulting in the following six image-based categories (**Figure 1**):

**Figure 1:**
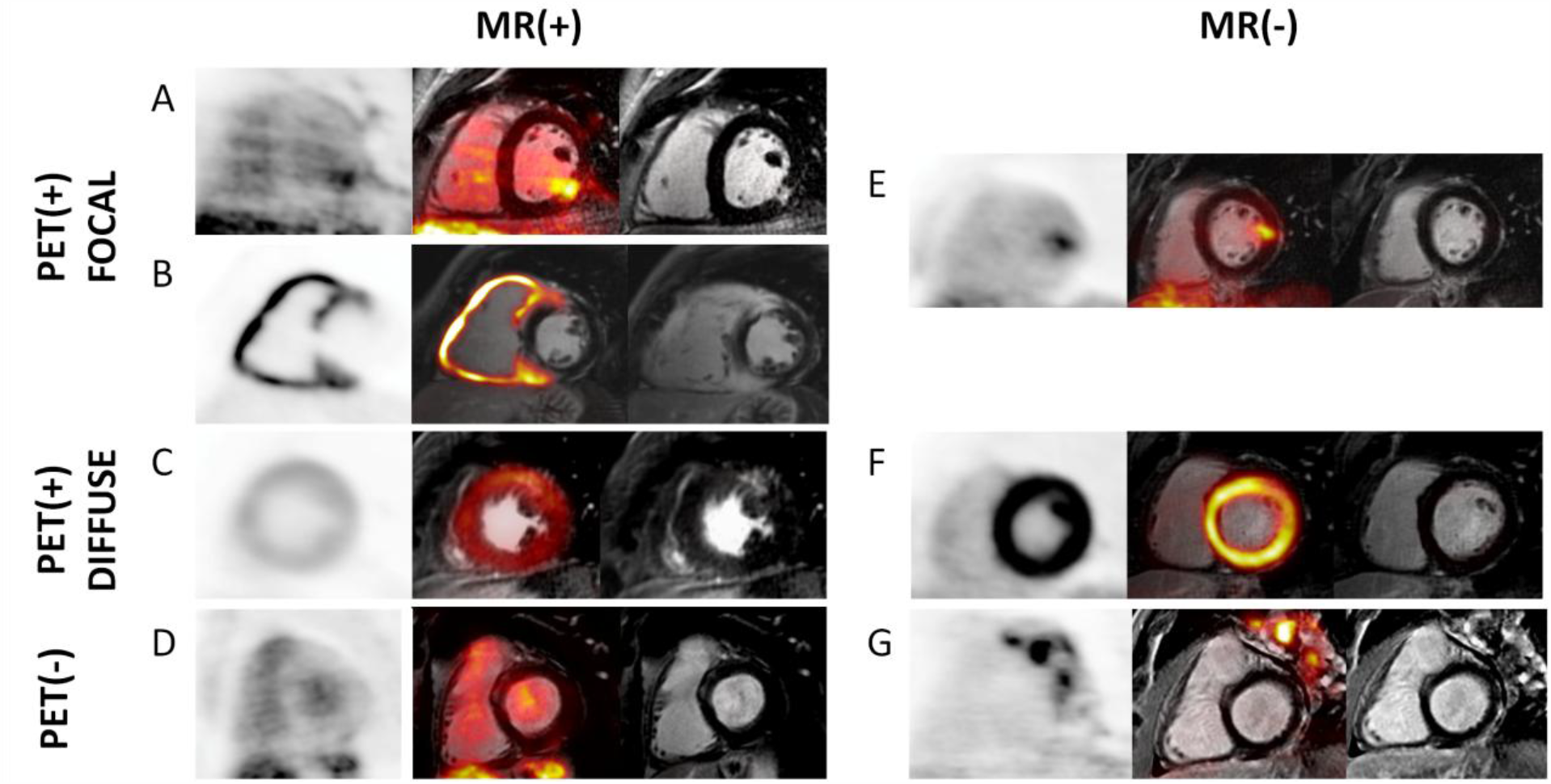
Combined PET/MR patterns. Representative short axis ^18^F-FDG PET (left), LGE MR (right) and fused MR/PET (middle) images for each of the six image-based categories. A: MR(+)PET(+)_FOCAL_, B: MR(+)PET(+)_FOCAL_ with predominantly right ventricular involvement that was subsequently confirmed by cardiac biopsy, C: MR(+)PET(+)_DIFFUSE_, D: MR(+)PET(-), E: MR(-)PET(+)_FOCAL_, F: MR(-)PET(+)_DIFFUSE_, G: MR(-)PET(-) with extra-cardiac ^18^F-FDG uptake.

1. MR(+)PET(+)_FOCAL_ when a characteristic pattern of LGE and a pattern of increased focal ^18^F-FDG uptake were both present – assigned as positive active CS [aCS(+)].
2. MR(+)PET(+)_DIFFUSE_ when a characteristic pattern of LGE was present and diffuse ^18^F-FDG uptake was present, suggestive of failed suppression of physiological uptake – assigned as negative active CS [aCS(-)].
3. MR(+)PET(-), defined by characteristic LGE but no increased ^18^F-FDG – representative of inactive cardiac sarcoidosis with residual scar, and assigned aCS(-).
4. MR(-)PET(+)_FOCAL_ if there was increased patchy, focal ^18^F-FDG uptake in the absence of underlying LGE – assigned aCS(-).
5. MR(-)PET(+)_DIFFUSE_ if there was increased diffuse ^18^F-FDG uptake in the absence of underlying LGE – assigned aCS(-).
6. MR(-)PET(-) if there was neither characteristic LGE nor increased ^18^F-FDG – assigned aCS(-).

### Statistical Analysis

Descriptive statistics were used to characterize the cohort. Continuous variables were described as mean ± standard deviation when normally distributed, otherwise median and inter-quartile range. Categorical variables were presented as n (%). Quantitative PET and MR values were compared pair-wise between each of the 6 image-based categories, and between selected groupings of the categories, using a two-sample two-sided t-test. Receiver operating characteristic curve analysis was performed and area under the curve reported to evaluate the ability of quantitative PET parameters (SUVmax, TBRmax, TNMRmax) and T2 to separate aCS(+) from aCS(-). The Youden index was used to determine the optimal threshold that differentiated aCS(+) from aCS(-) patients and sensitivity and specificity measures were calculated based on the optimal threshold. The association between the occurrence of a cardiac-related clinical outcome and the dichotomous classification of patients based on various groupings of the image-based categories was analyzed by calculating odds ratios. All hypothesis testing was conducted at the 0.05 two-sided significance level using R V3.5.3.

## Results

The study cohort that underwent MR/PET imaging included 148 patients. The characteristics of the cohort are summarized in **Table 1**. T2 measurements were not obtained in all patients due to frequent image artifacts arising from the use of bSSFP for T2 mapping (**Table 1**). Examples of the 6 imaging categories used are shown in **Figure 1**. One patient (**Figure 1, B**) presented with ventricular tachycardia, in the absence of extracardiac manifestation of sarcoidosis. The patient was classified as MR(+)PET(+)_FOCAL_ due to LGE throughout the right ventricular wall and at the right ventricular insertion points suggestive of CS, that coincided with widespread patchy/focal ^18^F-FDG uptake. The diagnosis of aCS(+) was subsequently confirmed by endomyocardial biopsy and the patient met the clinical endpoint, receiving an ICD.

**Table 1:**
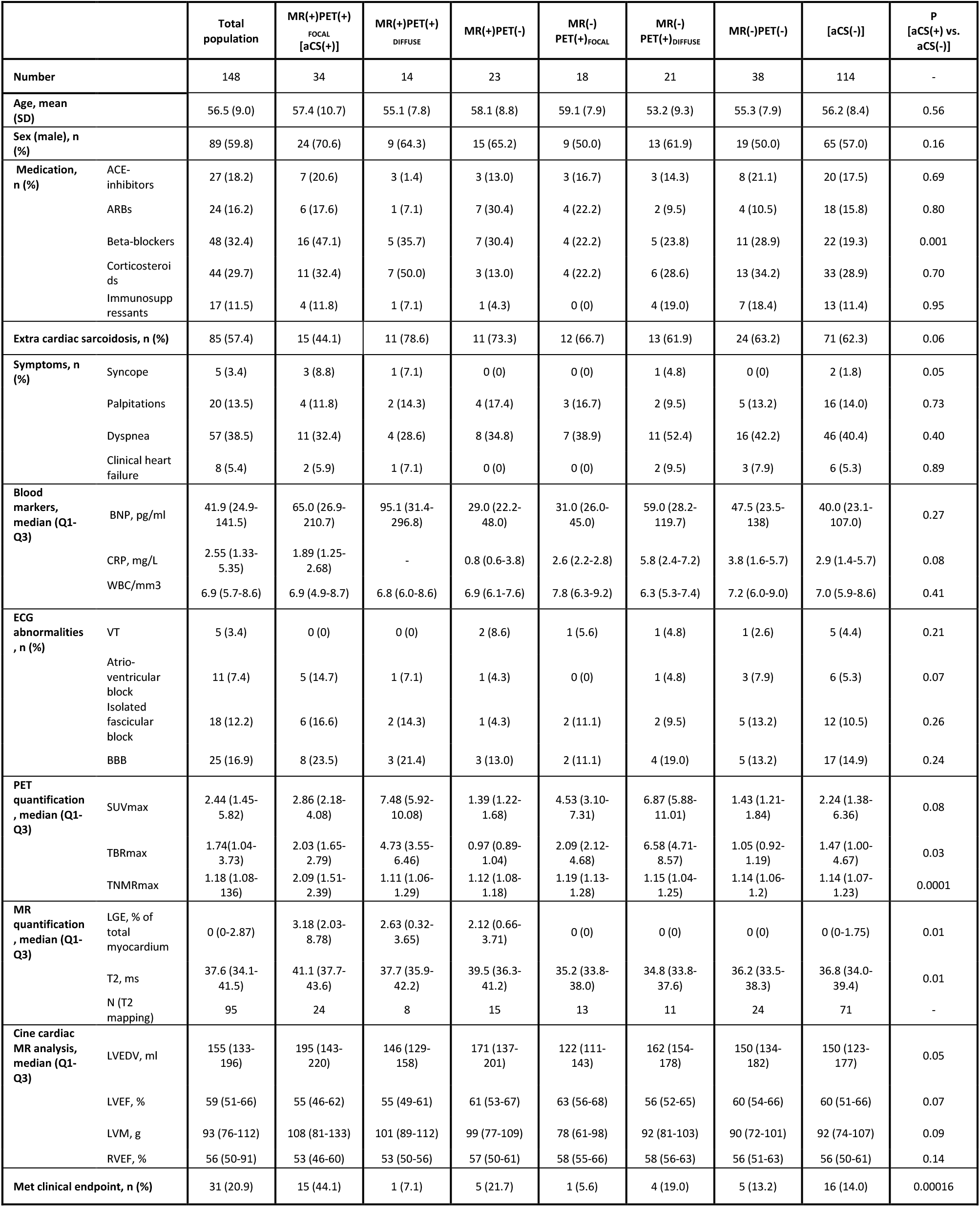
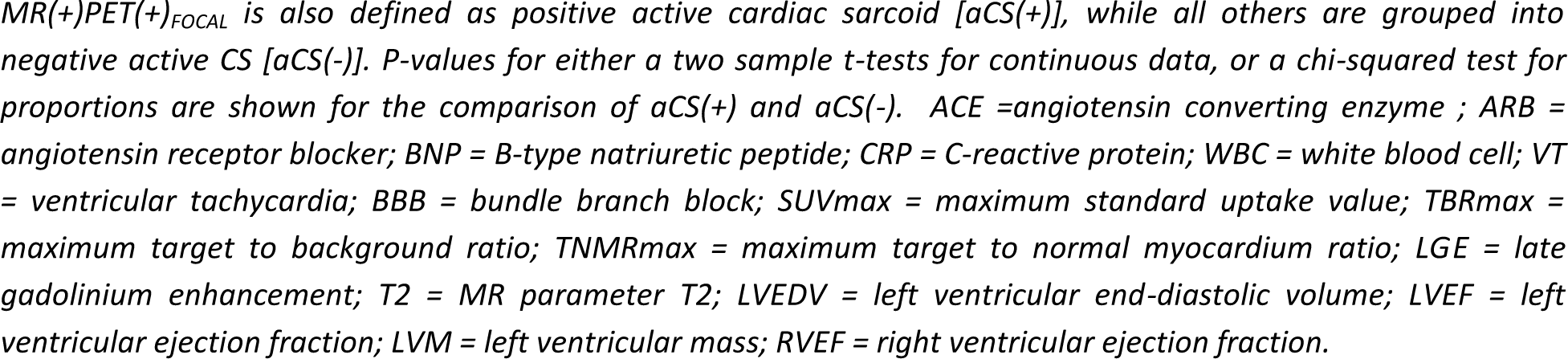
Clinical attributes of the cohort for each MR/PET imaging designation.

|  |  | Total population | MR(+)PET(+) FOCAL [aCS(+)] | MR(+)PET(+) DIFFUSE | MR(+)PET(-) | MR(-) PET(+)FOCAL | MR(-) PET(+)DIFFUSE | MR(-)PET(-) | [aCS(-)] | P [aCS(+) vs. aCS(-)] |
| --- | --- | --- | --- | --- | --- | --- | --- | --- | --- | --- |
| Number |  | 148 | 34 | 14 | 23 | 18 | 21 | 38 | 114 | - |
| Age, mean (SD) |  | 56.5 (9.0) | 57.4 (10.7) | 55.1 (7.8) | 58.1 (8.8) | 59.1 (7.9) | 53.2 (9.3) | 55.3 (7.9) | 56.2 (8.4) | 0.56 |
| Sex (male), n (%) |  | 89 (59.8) | 24 (70.6) | 9 (64.3) | 15 (65.2) | 9 (50.0) | 13 (61.9) | 19 (50.0) | 65 (57.0) | 0.16 |
| Medication, n (%) | ACE-inhibitors | 27 (18.2) | 7 (20.6) | 3 (1.4) | 3 (13.0) | 3 (16.7) | 3 (14.3) | 8 (21.1) | 20 (17.5) | 0.69 |
|  | ARBs | 24 (16.2) | 6 (17.6) | 1 (7.1) | 7 (30.4) | 4 (22.2) | 2 (9.5) | 4 (10.5) | 18 (15.8) | 0.80 |
|  | Beta-blockers | 48 (32.4) | 16 (47.1) | 5 (35.7) | 7 (30.4) | 4 (22.2) | 5 (23.8) | 11 (28.9) | 22 (19.3) | 0.001 |
|  | Corticosteroids | 44 (29.7) | 11 (32.4) | 7 (50.0) | 3 (13.0) | 4 (22.2) | 6 (28.6) | 13 (34.2) | 33 (28.9) | 0.70 |
|  | Immunosuppressants | 17 (11.5) | 4 (11.8) | 1 (7.1) | 1 (4.3) | 0 (0) | 4 (19.0) | 7 (18.4) | 13 (11.4) | 0.95 |
| Extra cardiac sarcoidosis, n (%) |  | 85 (57.4) | 15 (44.1) | 11 (78.6) | 11 (73.3) | 12 (66.7) | 13 (61.9) | 24 (63.2) | 71 (62.3) | 0.06 |
| Symptoms, n (%) | Syncope | 5 (3.4) | 3 (8.8) | 1 (7.1) | 0 (0) | 0 (0) | 1 (4.8) | 0 (0) | 2 (1.8) | 0.05 |
|  | Palpitations | 20 (13.5) | 4 (11.8) | 2 (14.3) | 4 (17.4) | 3 (16.7) | 2 (9.5) | 5 (13.2) | 16 (14.0) | 0.73 |
|  | Dyspnea | 57 (38.5) | 11 (32.4) | 4 (28.6) | 8 (34.8) | 7 (38.9) | 11 (52.4) | 16 (42.2) | 46 (40.4) | 0.40 |
|  | Clinical heart failure | 8 (5.4) | 2 (5.9) | 1 (7.1) | 0 (0) | 0 (0) | 2 (9.5) | 3 (7.9) | 6 (5.3) | 0.89 |
| Blood markers, median (Q1-Q3) | BNP, pg/ml | 41.9 (24.9-141.5) | 65.0 (26.9-210.7) | 95.1 (31.4-296.8) | 29.0 (22.2-48.0) | 31.0 (26.0-45.0) | 59.0 (28.2-119.7) | 47.5 (23.5-138) | 40.0 (23.1-107.0) | 0.27 |
|  | CRP, mg/L | 2.55 (1.33-5.35) | 1.89 (1.25-2.68) | - | 0.8 (0.6-3.8) | 2.6 (2.2-2.8) | 5.8 (2.4-7.2) | 3.8 (1.6-5.7) | 2.9 (1.4-5.7) | 0.08 |
|  | WBC/mm3 | 6.9 (5.7-8.6) | 6.9 (4.9-8.7) | 6.8 (6.0-8.6) | 6.9 (6.1-7.6) | 7.8 (6.3-9.2) | 6.3 (5.3-7.4) | 7.2 (6.0-9.0) | 7.0 (5.9-8.6) | 0.41 |
| ECG abnormalities, n (%) | VT | 5 (3.4) | 0 (0) | 0 (0) | 2 (8.6) | 1 (5.6) | 1 (4.8) | 1 (2.6) | 5 (4.4) | 0.21 |
|  | Atrio-ventricular block | 11 (7.4) | 5 (14.7) | 1 (7.1) | 1 (4.3) | 0 (0) | 1 (4.8) | 3 (7.9) | 6 (5.3) | 0.07 |
|  | Isolated fascicular block | 18 (12.2) | 6 (16.6) | 2 (14.3) | 1 (4.3) | 2 (11.1) | 2 (9.5) | 5 (13.2) | 12 (10.5) | 0.26 |
|  | BBB | 25 (16.9) | 8 (23.5) | 3 (21.4) | 3 (13.0) | 2 (11.1) | 4 (19.0) | 5 (13.2) | 17 (14.9) | 0.24 |
| PET quantification, median (Q1-Q3) | SUVmax | 2.44 (1.45-5.82) | 2.86 (2.18-4.08) | 7.48 (5.92-10.08) | 1.39 (1.22-1.68) | 4.53 (3.10-7.31) | 6.87 (5.88-11.01) | 1.43 (1.21-1.84) | 2.24 (1.38-6.36) | 0.08 |
|  | TBRmax | 1.74(1.04-3.73) | 2.03 (1.65-2.79) | 4.73 (3.55-6.46) | 0.97 (0.89-1.04) | 2.09 (2.12-4.68) | 6.58 (4.71-8.57) | 1.05 (0.92-1.19) | 1.47 (1.00-4.67) | 0.03 |
|  | TNMRmax | 1.18 (1.08-136) | 2.09 (1.51-2.39) | 1.11 (1.06-1.29) | 1.12 (1.08-1.18) | 1.19 (1.13-1.28) | 1.15 (1.04-1.25) | 1.14 (1.06-1.2) | 1.14 (1.07-1.23) | 0.0001 |
| MR quantification, median (Q1-Q3) | LGE, % of total myocardium | 0 (0-2.87) | 3.18 (2.03-8.78) | 2.63 (0.32-3.65) | 2.12 (0.66-3.71) | 0 (0) | 0 (0) | 0 (0) | 0 (0-1.75) | 0.01 |
|  | T2, ms | 37.6 (34.1-41.5) | 41.1 (37.7-43.6) | 37.7 (35.9-42.2) | 39.5 (36.3-41.2) | 35.2 (33.8-38.0) | 34.8 (33.8-37.6) | 36.2 (33.5-38.3) | 36.8 (34.0-39.4) | 0.01 |
|  | N (T2 mapping) | 95 | 24 | 8 | 15 | 13 | 11 | 24 | 71 | - |
| Cine cardiac MR analysis, median (Q1-Q3) | LVEDV, ml | 155 (133-196) | 195 (143-220) | 146 (129-158) | 171 (137-201) | 122 (111-143) | 162 (154-178) | 150 (134-182) | 150 (123-177) | 0.05 |
|  | LVEF, % | 59 (51-66) | 55 (46-62) | 55 (49-61) | 61 (53-67) | 63 (56-68) | 56 (52-65) | 60 (54-66) | 60 (51-66) | 0.07 |
|  | LVM, g | 93 (76-112) | 108 (81-133) | 101 (89-112) | 99 (77-109) | 78 (61-98) | 92 (81-103) | 90 (72-101) | 92 (74-107) | 0.09 |
|  | RVEF, % | 56 (50-91) | 53 (46-60) | 53 (50-56) | 57 (50-61) | 58 (55-66) | 58 (56-63) | 56 (51-63) | 56 (50-61) | 0.14 |
| Met clinical endpoint, n (%) |  | 31 (20.9) | 15 (44.1) | 1 (7.1) | 5 (21.7) | 1 (5.6) | 4 (19.0) | 5 (13.2) | 16 (14.0) | 0.00016 |

$MR(+)/PET(+)_\text{FOCAL}$ is also defined as positive active cardiac sarcoid [aCS(+)], while all others are grouped into negative active CS [aCS(-)]. P-values for either a two sample t-tests for continuous data, or a chi-squared test for proportions are shown for the comparison of aCS(+) and aCS(-). ACE = angiotensin converting enzyme ; ARB = angiotensin receptor blocker; BNP = B-type natriuretic peptide; CRP = C-reactive protein; WBC = white blood cell; VT = ventricular tachycardia; BBB = bundle branch block; SUVmax = maximum standard uptake value; TBRmax = maximum target to background ratio; TNMRmax = maximum target to normal myocardium ratio; LGE = late gadolinium enhancement; T2 = MR parameter T2; LVEDV = left ventricular end-diastolic volume; LVEF = left ventricular ejection fraction; LVM = left ventricular mass; RVEF = right ventricular ejection fraction.

Analysis of the quantitative PET parameters, by pair-wise comparison between each image-based category, indicated that TNMRmax was significantly higher in the aCS(+) group [MR(+)PET(+)_FOCAL_] than all others (**Figure 2** and **Table 2**). SUVmax and TBRmax were significantly higher for aCS(+) than any PET(-) category and lower than the remaining PET(+) categories. Similarly, T2 values for aCS(+) were significantly higher than any MR(-) category but not significantly different from the other MR(+) categories, suggesting T2 values are affected by both the edematous component of inflammation(16) and myocardial scarring. Receiver operating characteristic curve analysis indicated that TNMRmax was an excellent predictor of the image-based designation of aCS(+), with an area under the curve (AUC) of 0.90 (**Figure 3**). The sensitivity and specificity were 88% and 95% at the Youden index point with a TNMRmax threshold for separating aCS(+) from aCS(-) of 1.4. For quantitative T2 measurements, the values of AUC/sensitivity/specificity were 0.72/79%/58% with a threshold of 37 ms. SUVmax and TBRmax were poor determinants of aCS(+), with AUCs of 0.54 and 0.57 (**Table 3**).

**Table 2:** Group-by-group comparison of two-sample two-sided t-tests for quantitative PET and MR parameters.

|  | MR(+)<br>PET(+)<sub>DIFFUSE</sub> |  |  |  | MR(+)<br>PET(-) |  |  |  | MR(-)<br>PET(+)<sub>FOCAL</sub> |  |  |  | MR(-)<br>PET(+)<sub>DIFFUSE</sub> |  |  |  | MR(-)<br>PET(-) |  |  |  |
| --- | --- | --- | --- | --- | --- | --- | --- | --- | --- | --- | --- | --- | --- | --- | --- | --- | --- | --- | --- | --- |
|  | SUV | TBR | TNMR | T2 | SUV | TBR | TNMR | T2 | SUV | TBR | TNMR | T2 | SUV | TBR | TNMR | T2 | SUV | TBR | TNMR | T2 |
| MR(+)<br>PET(+)<sub>FOCAL</sub> | ** | ** | ** | ns | ** | ** | ** | ns | * | * | ** | * | ** | ** | ** | ** | ** | ** | ** | ** |
| MR(+)<br>PET(+)<sub>DIFFUSE</sub> |  |  |  |  | ** | ** | ns | ns | ns | ns | ns | ns | ns | ns | ns | ns | ** | ** | ns | ns |
| MR(+)<br>PET(-) |  |  |  |  |  |  |  |  | ** | ** | ns | ns | ** | ** | ns | * | ns | * | ns | * |
| MR(-)<br>PET(+)<sub>FOCAL</sub> |  |  |  |  |  |  |  |  |  |  |  |  | * | ** | ns | ns | ** | ** | ns | ns |
| MR(-)<br>PET(+)<sub>DIFFUSE</sub> |  |  |  |  |  |  |  |  |  |  |  |  |  |  |  |  | ** | ** | ns | ns |

*SUVmax* = maximum standard uptake value, *TBRmax* = maximum target to background ratio, *TNMRmax* = maximum target to normal myocardium ratio, and *T2* = MR parameter T2. \*\* = $p < 0.01$ , \* = $p < 0.05$ , ns = not significant.

**Table 3:**
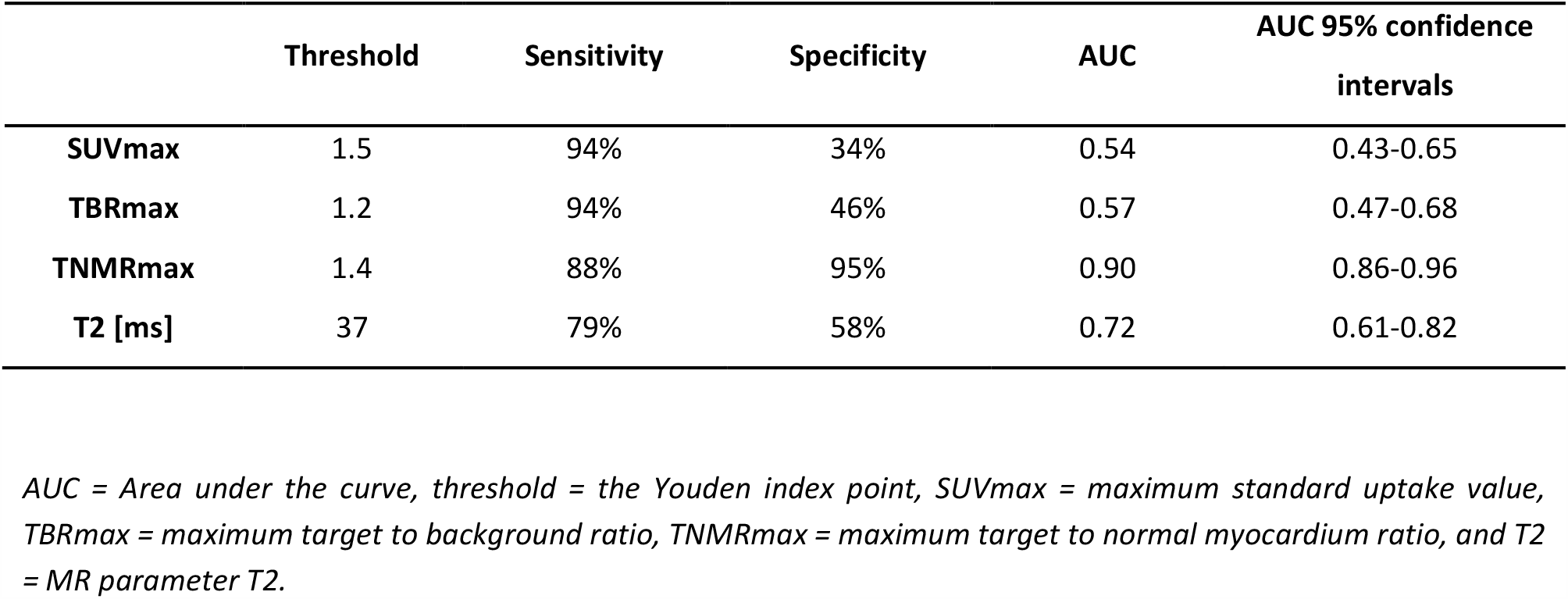
Receiver operating characteristic curve analysis for separating aCS(+) [MR(+)PET(+)_FOCAL_] from aCS(-) [all others].

**Figure 2:**
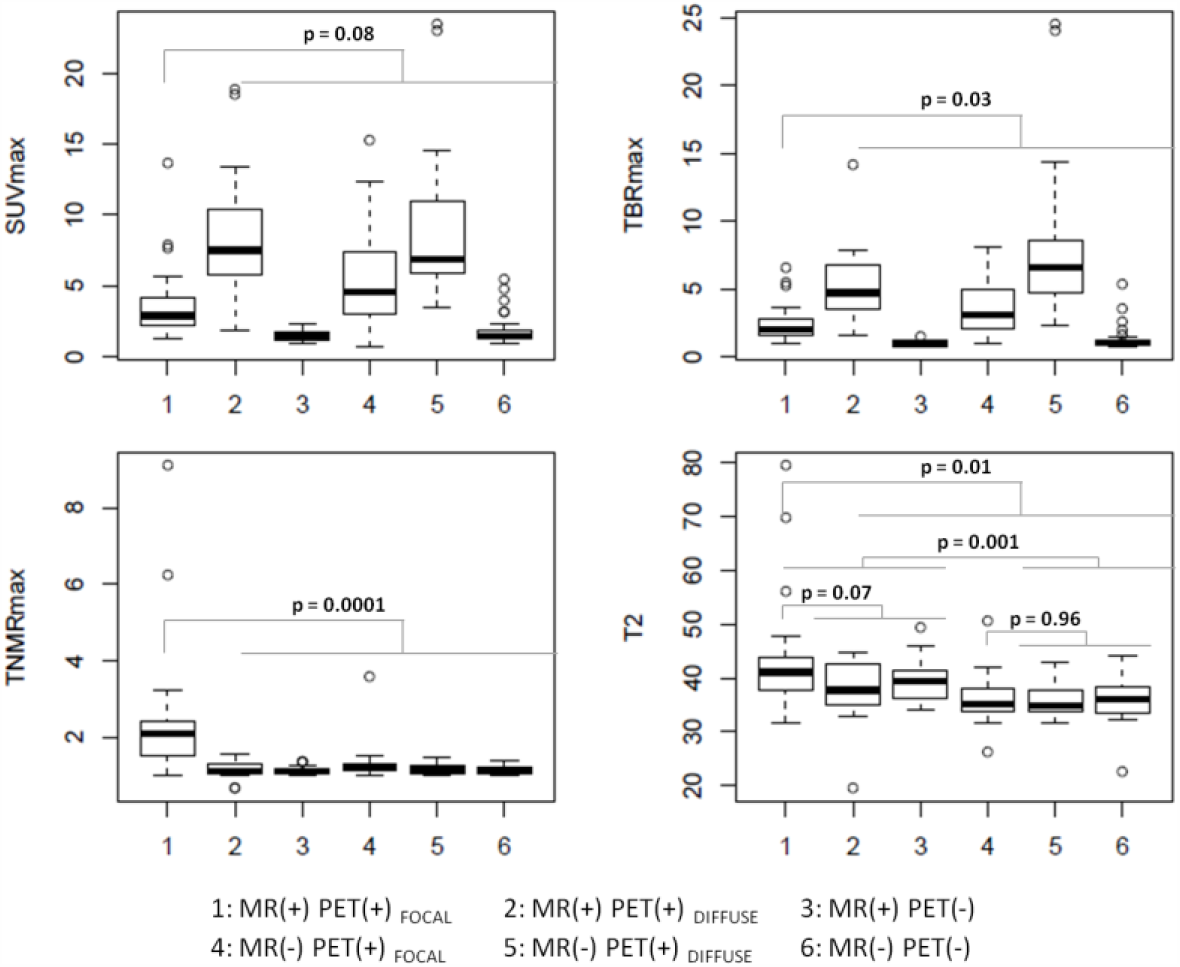
Quantitative evaluation of PET and T2 parameters for each image-based category. Box-and-whisker plots for the quantitative PET and T2 parameters for each image-based category. P-values are from two-sided two-sample t-tests between selected groupings of image-based categories.

**Figure 3:**
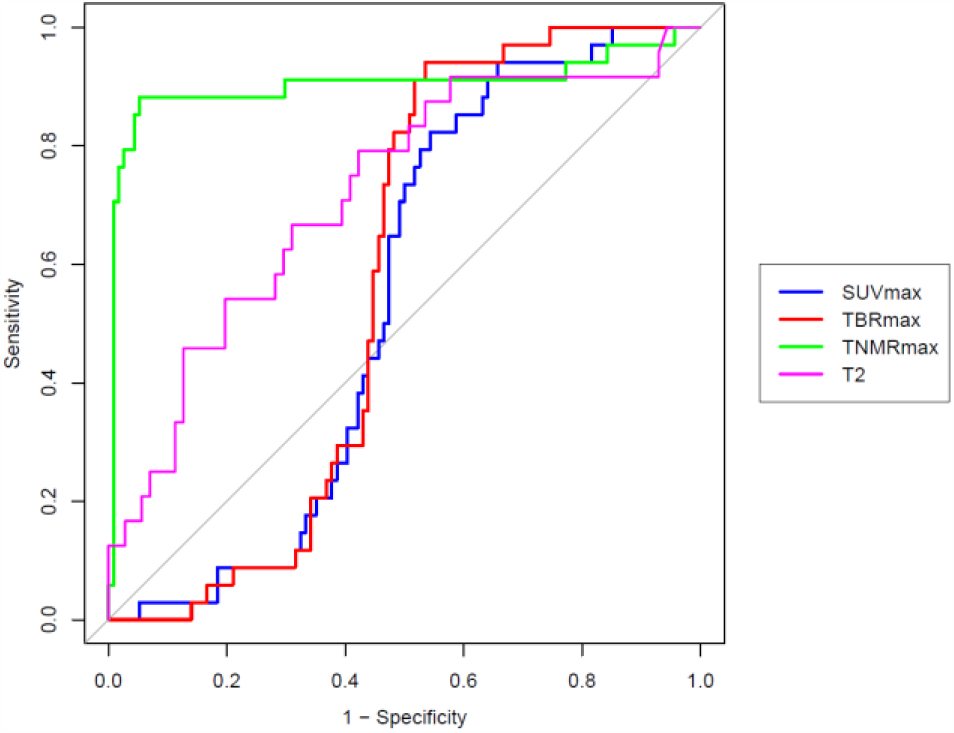
Receiver operating characteristic curve analysis. Receiver operating characteristic curve analysis for separating aCS(+) [MR(+)PET(+)_FOCAL_] from aCS(-) [all other image-based categories].

Thirty one (21%) of the 148 patients reached the composite clinical endpoint (**Table 4**). Of those, all had a cardiac device implanted. The proportions of patients in the aCS(+) and aCS(-) groups that reached the clinical endpoint, as well as other groupings of the 6 imaging categories, are given in **Table 5**. The unadjusted odds ratios (OR) and confidence intervals (CI) resulting from those groupings are given in **Table 6**, showing that patients with an imaging designation of aCS(+) had 4.8 times higher odds of reaching the clinical outcome compared to aCS(-) patients (OR 4.8; 95% CI 2.0-11.4; p<0.001). When the analysis was conducted without accounting for the pattern of ^18^F-FDG uptake, MR(+)PET(+) cases [combining MR(+)PET(+)_FOCAL_ and MR(+)PET(+)_DIFFUSE_] vs. all others combined, showed a similarly significant association with clinical outcomes (OR 2.8; 95% CI 1.3-6.4; p=0.012) as MR(+) vs. MR(-) (OR 2.8; 95% CI 1.2-6.5; p=0.015). PET(+) alone [PET(+)_FOCAL_ and PET(+)_DIFFUSE_ combined vs. PET(-)] showed a trend towards being associated with the clinical outcome, which did not reach statistical significance (OR 1.6; 95% CI 0.7-3.7; p=0.26).

**Table 4:** Patients meeting the composite clinical endpoint.

| Endpoint | MR(+) PET(+)<sub>FOCAL</sub> | MR(+) PET(+)<sub>DIFFUSE</sub> | MR(+) PET(-) | MR(-) PET(+)<sub>FOCAL</sub> | MR(-) PET(+)<sub>DIFFUSE</sub> | MR(-) PET(-) | Total |
| --- | --- | --- | --- | --- | --- | --- | --- |
| **ICD and VT and aborted cardiac arrest** | 1 (3%) | - | - | - | - | - | 1 (3%) |
| **ICD and VT** | 1 (3%) | - | 1 (3%) | - | - | - | 2 (6%) |
| **ICD and CHB** | 1 (3%) | - | - | - | - | - | 1 (3%) |
| **ICD** | 3 (10%) | - | - | 1 (3%) | 2 (6%) | 1 (3%) | 7 (23%) |
| **S-ICD** | - | - | - | - | 1 (3%) | 1 (3%) | 2 (6%) |
| **WCD** | - | - | - | - | 1 (3%) | - | 1 (3%) |
| **CRT-D** | 3 (10%) | - | - | - | - | - | 3 (10%) |
| **ILR** | 6 (19%) | 1 (3%) | 4 (13%) | - | - | 3 (10%) | 14 (45%) |
| **Total** | 15 (48%) | 1 (3%) | 5 (16%) | 1 (3%) | 4 (13%) | 5 (16%) | 31 (100%) |

*Number of patients and percent of all patients meeting the clinical endpoint (in parentheses) are given in the table. ICD = implantable cardioverter defibrillator; VT = ventricular tachycardia; CHB = complete heart block; S-ICD = subcutaneous implantable cardioverter defibrillator; WCD = wearable cardioverter defibrillator; CRT-D = cardiac resynchronization therapy device; ILR = implantable loop recorder.*

**Table 5:** Cardiac-related clinical outcome events by analysis groups for each group.

| Analysis |  | Event | Non-event | Events-to-non-events ratio (%) |
| --- | --- | --- | --- | --- |
| **1** | aCS(+) | 15 (10) | 19 (13) | 79 |
|  | aCS(-) | 16 (11) | 98 (66) | 16 |
| **2** | MR(+)/PET(+) | 16 (11) | 32 (22) | 50 |
|  | Non MR(+)/PET(+) | 15 (10) | 85 (57) | 18 |
| **3** | PET(+) | 21 (14) | 66 (45) | 32 |
|  | PET(-) | 10 (7) | 51 (34) | 20 |
| **4** | MR(+) | 21 (14) | 50 (34) | 42 |
|  | MR(-) | 10 (7) | 67 (45) | 15 |

*Events and non-events are given as number (n) and percent of all patients (%) in parentheses. Analysis 1: aCS(+) [MR(+)/PET(+)<sub>FOCAL</sub>] vs. aCS(-) [all others]; analysis 2: MR(+)/PET(+) [MR(+)/PET(+)<sub>FOCAL</sub> and MR(+)/PET(+)<sub>DIFFUSE</sub>] vs. Non MR(+)/PET(+) [all others]; analysis 3: PET(+) [MR(+)/PET(+)<sub>FOCAL</sub>, MR(-)/PET(+)<sub>FOCAL</sub>, MR(+)/PET(+)<sub>DIFFUSE</sub>, and MR(-)/PET(+)<sub>DIFFUSE</sub>] vs. PET(-) [all others]; analysis 4: MR(+) [MR(+)/PET(+)<sub>FOCAL</sub>, MR(+)/PET(+)<sub>DIFFUSE</sub>, MR(+)/PET(-)] vs. MR(-) [all others].*

**Table 6:** Association of analysis groups with cardiac-related clinical outcomes.

| Analysis | Odds of having a cardiac-related clinical outcome/event | OR | CI | P |
| --- | --- | --- | --- | --- |
| 1 | aCS(+) vs. aCS(-) | 4.8 | 2.0 – 11.4 | 0.00032 |
| 2 | MR(+)PET(+) vs. non MR(+)PET(+) | 2.8 | 1.3 – 6.4 | 0.012 |
| 3 | PET(+) vs. PET(-) | 1.6 | 0.7 – 3.7 | 0.26 |
| 4 | MR(+) vs. MR(-) | 2.8 | 1.2 – 6.5 | 0.015 |

*Analysis 1: aCS(+) [MR(+)PET(+)<sub>FOCAL</sub>] vs. aCS(-) [all others]; analysis 2: MR(+)PET(+) [MR(+)PET(+)<sub>FOCAL</sub> and MR(+)PET(+)<sub>DIFFUSE</sub>] vs. Non MR(+)PET(+) [all others]; analysis 3: PET(+) [MR(+)PET(+)<sub>FOCAL</sub>, MR(-)PET(+)<sub>FOCAL</sub>, MR(+)PET(+)<sub>DIFFUSE</sub>, and MR(-)PET(+)<sub>DIFFUSE</sub>] vs. PET(-) [all others]; analysis 4: MR(+) [MR(+)PET(+)<sub>FOCAL</sub>, MR(+)PET(+)<sub>DIFFUSE</sub>, MR(+)PET(-)] vs. MR(-) [all others]. OR = odds ratio; CI = 95% confidence interval; p-values are for a two-sided test of significance for the odds ratio.*

## Discussion

This study, with a substantially larger cohort than previous studies of MR/PET and CS, has further validated the use of MR/PET in the evaluation of CS, and specifically of TNMRmax(9) as an effective discriminator of positive active CS (**Figure 2**). Importantly, we also show a significant association between positive active CS and outcomes, with patients categorized as aCS(+) on MR/PET demonstrating more than 4-fold higher odds for progressing to a cardiac-related event and/or implant of a cardiac device (**Table 6**). The significant associations found in this study suggest that MR/PET may be able to predict future clinical events and provide useful prognostic information. Understanding disease course and prognosis with MR/PET represents a significant step and suggests that this imaging technique might be useful in guiding the initiation and monitoring of suitable anti-inflammatory or immune suppressant therapies that have shown potential benefit in CS.(17,18) In addition, the association of the imaging definition of aCS(+) and clinical events validates our strict diagnostic approach to the detection of active CS – which requires both positive PET and MR findings, and excludes more diffuse patterns of FDG-PET; it also suggests that MR/PET alone might be a useful alternative to invasive endomyocardial biopsy. The higher use of beta blockers, prevalence of syncope and the trend for increased prevalence of atrioventricular block in aCS(+) patients (**Table 1**) further corroborates our finding that classification of aCS(+) by imaging is consistent with disease.

In this study we expanded the image-based categories from 4 to 6 by subdividing the PET(+) categories based on the qualitative appearance and pattern of tracer uptake (**Figure 1**). PET(+)_FOCAL_ described ^18^F-FDG uptake in a patchy/focal distribution consistent with the guideline’s description of CS, and PET(+)_DIFFUSE_ described diffuse ^18^F-FDG uptake consistent with physiological uptake due to failed myocardial suppression. These additional categories may increase the specificity for the imaging designation of positive active CS by eliminating false positive failed myocardial suppression with quiescent disease [MR(+)PET(+)_DIFFUSE_] from the aCS(+) group. Without categorization by pattern of ^18^F-FDG uptake, these cases (especially for patients with extra-cardiac disease and strong suspicion of CS) would otherwise be categorized as MR(+)PET(+), and consequently aCS(+). This sub-categorization is supported by our finding that MR(+)PET(+)_FOCAL_ vs. all others showed the highest odds ratio associating the imaging categorization with a cardiac-related clinical outcome, higher than MR(+)PET(+) alone [MR(+)PET(+)_FOCAL_ and MR(+)PET(+)_DIFFUSE_ vs. all others] (**Table 6**). Nevertheless, for subjects classified as MR(+)PET(+)_DIFFUSE_, herein designated as aCS(-), it is important to emphasize that active CS could not be completely ruled out since subtle, “true” focal or focal-on-diffuse CS activity might have been masked by background physiologic ^18^F-FDG uptake. Furthermore, it is conceivable that the additional imaging category, MR(-)PET(+)_FOCAL_, which we classified as aCS(-), might possibly have greater sensitivity for early inflammatory changes and allow identification of pathological uptake prior to the development of myocardial injury as has been suggested by other investigators(19). Due to the small group sizes, the lack of histological verification or longer follow-up to evaluate for progression from MR(-)PET(+)_FOCAL_ to MR(+)PET(+)_FOCAL_, the pathophysiological correlates of this particular imaging category were not investigated. Further research will be required to better categorize the MR(+)PET(+)_DIFFUSE_ and MR(-)PET(+)_FOCAL_ subgroups and to determine when and if serial, interval imaging or close monitoring for disease progression, especially for cases with a high clinical suspicion of active disease, is warranted. Taken together, these finding suggest that the combined interpretation of PET and MR data coupled with the patchy/focal pattern of the ^18^F-FDG uptake, is a better indicator of association with a cardiac-related clinical outcome and, therefore, disease severity.

In addition to the qualitative metrics described above, quantitative values of SUVmax and TBRmax in the aCS(+) group [MR(+)PET(+)_FOCAL_] were significantly lower than other PET(+) categories [MR(+)PET(+)_DIFFUSE_, MR(-)PET(+)_FOCAL_, MR(-)PET(+)_DIFFUSE_]. These data indicate that quantitative analysis of SUVmax and TBRmax may allow discrimination between uptake associated with inflammation vs. physiological or non-specific uptake.

Together with the pattern of ^18^F-FDG uptake, analysis of T2 values (**Figure 2**) may prove useful in separating pathological from physiological uptake. When all MR(+) cases were combined and compared to all MR(-) cases combined, T2 for the MR(+) group (41.0 ± 9.1 ms) was significantly higher than for the MR(-) group (36.1 ± 4.6 ms), p=0.001, correlating with the presence of LGE. Notably, within the MR(+) groups, there was a trend (p=0.07) for MR(+)PET(+)_FOCAL_ to show higher T2 values (43.3 ± 11.0 ms) compared to MR(+)PET(+)_DIFFUSE_ and MR(+)PET(-) groups combined (38.6 ± 5.9 ms). These data indicate an association between T2 and focal ^18^F-FDG uptake characteristic of inflammation that occurs in the presence of LGE. For the MR(-) groups, there was no comparable significant difference (p=0.96) between the MR(-)PET(+)_FOCAL_ category (36.1 ± 5.7 ms) and the MR(-)PET(+)_DIFFUSE_ and MR(-)PET(-) groups combined (36.0 ± 4.2 ms).

The expansion of the imaging categories coupled with the T2 analysis suggest the potential utility of more sophisticated multi-parametric models to characterize active CS disease and separate pathological, physiological and mixed ^18^F-FDG uptake. A future model would include analysis of TNMRmax to characterize patchy/focal ^18^F-FDG uptake, SUVmax to characterize physiological ^18^F-FDG uptake, as well as the MR parameters: LGE, to define myocardial injury, and T2, to confirm myocardial inflammation. In addition, secondary parameters such as spatial heterogeneity of ^18^F-FDG uptake(20) and kinetic analysis of ^18^F-FDG uptake(9,21) could be useful. Such models would require histological validation.

### Limitations

The evaluation of prognosis is limited by the relatively small patient cohort and the limited number who progressed to the clinical endpoint within the follow-up period. Furthermore, a relatively short duration of follow-up in this cohort study has limited the extent of clinical progression that would be necessary to provide a more detailed evaluation of the prognostic ability of the imaging results. Further studies with larger cohorts, longer follow-up and time-to-event models are required to evaluate clinical manifestations specific to prognosis of CS, beyond the cardiac-related clinical outcomes used in this study. Device implantation, which was ubiquitous in those who met the clinical endpoint, is related to arrhythmia and therefore was, *a priori*, expected to correlate with scarring seen on LGE MR.(22,23) The follow-up period was not sufficient to evaluate the impact of early stages of inflammation that could be reflected by MR(-)PET(+) findings, to manifest in scarring, arrhythmias and device implantation, therefore the impact of the PET(+) imaging designation is likely underestimated, consistent with the fact that MR(+) alone [MR(+)PET(+)_FOCAL,_ MR(+)PET(+)_DIFFUSE_ and MR(+)PET(-) vs. all others] showed a higher odds of progressing to the clinical outcome than PET(+) alone [MR(+)PET(+)_FOCAL,_ MR(+)PET(+)_DIFFUSE_, MR(-)PET(+)_FOCAL_, and MR(-)PET(+)_DIFFUSE_ vs. all others]. An additional limitation is the lack of a gold standard confirmation that inflammation is the underlying mechanism of myocardial ^18^F-FDG uptake.

^18^F-FDG has been correlated with inflammation in other applications(24) yet we cannot rule-out increased metabolic activity of the cardiomyocytes as the underlying mechanism for the increased myocardial ^18^F-FDG uptake observed in the setting of CS. Use of novel tracers to assess myocardial inflammation without background myocardial signal such as ^68^Ga-DOTATATE(25) or ^68^Ga-DOTANOC(26) may be useful to confirm the findings.

## Conclusions

Hybrid MR/PET imaging with an extended image-based classification for diagnosis of CS is associated with cardiac-related clinical outcomes in CS. Moreover, target-to-normal-myocardium ratio (TNMRmax) has high sensitivity and excellent specificity for quantifying the imaging-based classification of active CS. Further studies are required to validate the imaging-based strategy for diagnosis and prognosis.

## Data Availability

All data produced in the present study are available upon reasonable request to the authors

## List of abbreviations

CS: Cardiac Sarcoidosis
^18^F-FDG: ^18^F-fluorodeoxyglucose
PET: Positron Emission Tomography
MR: Magnetic Resonance
LGE: Late Gadolinium Enhancement
SUVmax: Maximum Standard Uptake Value
TBRmax: Maximum Target-to-Background Ratio
TNMRmax: Maximum Target-to-Normal-Myocardium Ratio
aCS(+): positive for active Cardiac Sarcoidosis
aCS(-): negative for active Cardiac Sarcoidosis
CRT-D: cardiac resynchronization therapy device
ICD: implantable cardioverter defibrillator
S-ICD: subcutaneous implantable cardioverter defibrillator
WCD: wearable cardioverter defibrillator
ILR: implantable loop recorder
bSSFP: balanced Steady-State Free-Precession
LV: Left Ventricle
RV: Right Ventricle
AUC: Area Under the Curve
OR: Odds Ratio
CI: Confidence Interval

